# The Training needs of registered midwives in two Comprehensive Emergency Obstetrics and Neonatal Care facilities in Eswatini

**DOI:** 10.64898/2026.03.13.26348361

**Authors:** Ntombifuthi J. Gama, Roinah Ngunyulu

**Affiliations:** Department of Midwifery, University of Eswatini, Mbabane, Kingdom of Eswatini; Department of Nursing, University of Johannesburg, Johannesburg, South Africa

**Keywords:** Registered midwives, training needs, knowledge and skills, emergency obstetrics, newborn care

## Abstract

**Background:** Emergency obstetrics and newborn care training improves the knowledge and skills of healthcare professionals. However, there is limited evidence on training programs that had been informed by training needs analysis. The aim of the study was to determine training needs of registered midwives to inform a training program.

**Methods:** A descriptive cross-sectional study design was used to collect data from N=202 midwives who worked at two comprehensive emergency obstetrics and neonatal care hospitals. Simple random sampling was used to select respondents. Ethics approval was obtained before conducting the study. Data were collected from November 2023 to January 2024 using an adapted self-administered Hennessy Hicks Training Needs Questionnaire. SPSS version 29 was used to analyze the data. Descriptive statistics, means and standard deviations were calculated. The differences between task importance and task performance were determined for each of the measured items. A paired sample t-test was used to establish the significance of the differences between each of the five category pairs with p=<.05.

**Results:** The mean age of the 202 respondents was 38.06±6.9 years. The midwives predominantly fell into the age group 40–44 years (n=53, 22.2%), and they had an average of 5–9 years of work experience (n=75, 37.1%). Training needs were perceived for all the measured items. The research/ audit category emerged as the highest (M=2.23±1.05) training need, followed by clinical (1.94±0.55), administrative (1.70±1.03), communication (1.57±0.79) and supervisory tasks (1.14±0.76). Differences between each of the five category pairs were statistically significant with p=<.05. The highest specific training need was newborn resuscitation (n= 61, 30.2%).

**Conclusion:** The study highlights the need for training on research and clinical tasks.

**Recommendation:** Tailor training according to the identified needs for the effective management of emergency obstetrics and newborn complications.

## Introduction

Eswatini is a low- to middle-income country (LMIC) with a population of 1 230 506 by 2023 (1). Maternal and newborn mortalities remain a challenge ten years after adoption of the Sustainable Development Goals (SDGs) agenda. A report on progress with SDGs show that the maternal mortality ratio (MMR) in LMICs is 346 per 100 000 live births compared to 10 per 100 000 live births in high-income countries (2). In Eswatini, the MMR is 452 per 100 000 live births (3) and the neonatal mortality ratio is 21 per 1 000 live births (4) despite a skilled birth attendant being present in 93.4% of cases and institutional delivery of 92.7 % (4).

Globally, haemorrhage remains the leading cause of maternal mortalities, followed by hypertensive disorders of pregnancy, perinatal sepsis, and complications of abortions (5). Indirect obstetric causes have also been mentioned as a leading cause of maternal mortalities (6). On the other hand, newborn mortalities occur as a result of prematurity complications, intrapartum-related complications, sepsis, congenital abnormalities, and pneumonia (7). A majority of the maternal deaths are preventable (8) through nine life-saving interventions known as signal functions.

The life-saving interventions include the administration of parenteral antibiotics, parenteral uterotonics and parenteral anticonvulsants, manual removal of a retained placenta, the removal of retained products of conception, performing assisted vaginal delivery, neonatal resuscitation with bag and mask, blood transfusion, and caesarean section (9). In order to perform the signal functions competently, midwives require knowledge and skills with respect to emergency obstetrics and newborn care (EmONC).

The International Confederation of Midwives (ICM) prescribes competencies for midwifery practice across five categories. These include cross-functional competencies, competencies for sexual and reproductive health and rights (SRHR), competencies for antenatal care, competencies for care during labor and birth, and competencies for ongoing postnatal care (10). In addition to the competencies, the reduction of mortalities also requires adequate staffing and skilled teams to work in enabling environments (11). Support through infrastructure and professional and systems integration enables practice across the full scope of midwifery practice (12). Conditions that minimize the application of clinical skills affect the use of the skills. Infrequent use of clinical skills results in a skills decay (13). Correction of skills decay requires refresher training (14).

Training should be informed by a training needs analysis (TNA) to be effective. A TNA is systematic assessment of the gaps in knowledge, skills and attitudes of employees that can be addressed by training (15). Training needs analyses have been conducted widely in healthcare. Thirty-three articles from 18 countries with more than two-thirds conducted in high-income countries were identified by 2021, and all the studies used the Hennessy Hicks Questionnaire, either in English or translated to a local language (16). Adaptation of the tool according to the specified guidelines, which state that eight items can be swapped and 10 added, does not compromise its validity and reliability (17).

Where the Hennessy Hicks tool have been used, the research/ audit category often rated highest of the five categories (18–22). Variation is noted in the remaining categories. Training needs related to research and clinical tasks were identified in a cross-sectional study on snakebite management among midwives and doctors (18). For primary care nurses, research and child mental health needs have been identified (19). Among physicians, research and supervisory skills have been rated the highest (23). The composition of respondents was found to influence training needs results. The training needs of caregivers from five countries were basic nursing skills, specialization, and psychology-related tasks (24).

In a majority of TNA studies, external validity was compromised because of the use of a convenience sampling technique, which limits generalizability of results to other contexts. The results also aligned with the measured items adapted by the researchers, which varied widely. Therefore, a TNA had to be conducted for the midwives from two comprehensive emergency obstetrics and newborn care (CEmONC) facilities in Eswatini to identify specific training needs.

In Eswatini, health service delivery is stratified into five levels. Community healthcare lies at the base of the pyramid and the national referral hospital is at the highest point (25). In the middle of the pyramid are the clinics, public health units, health centers and regional referral hospitals. Midwives are the majority group of professionals who attend maternal and newborn healthcare at all levels of care. In most cases of perinatal complications and newborns with complications, midwives are the point of contact including after normal operating hours due to shortages of doctors and obstetricians. In an effort to sharpen the knowledge and skills of midwives with respect to the management of emergency and newborn complications, the Sexual and Reproductive Health Unit (SRHU) provides training across the country. Despite the training, mortalities remain high. It has therefore become necessary to determine training needs of registered midwives so that the training can respond to identified performance needs.

## Materials and methods

### Study sites

The study was conducted at two CEmONC hospitals in Eswatini. Study site A is a regional referral hospital situated in the central part of Eswatini in Manzini. Study site B is situated in Mbabane, the capital city of Eswatini and is a national referral hospital. Functionally, the national referral hospital has a neonatal intensive care unit (NICU). Otherwise, all other services are similar. Both hospitals perform CEmONC signal functions and operate for 24 hours a day, every day. Registered midwives, doctors, specialists, nursing assistants and allied health workers manage clients in these hospitals. Registered midwives were the target population for this study.

### Study design

A quantitative, descriptive, cross-sectional study design was used to determine the training needs of registered midwives at both study sites.

### Sampling technique and sample size

A simple random probability sampling method was used to select the sample because the sample was homogenous. A sampling frame of nurse midwives from each study site was drawn, taken from scheduling timetables. A random number table was generated and numbers that were picked from the table were matched with the sampling frame. To determine the sample size, the sample size calculation (26) was:

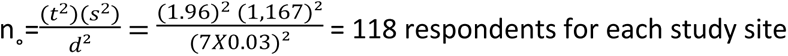

t=alpha level, 1.96

s= estimated standard deviation of 1,167 obtained by dividing the seven Likert scale responses over six possible standard deviations

d=acceptable margin of error, 0.03 in continuous outcome variable-Likert scale

### Data collection instrument

The Hennessy Hicks Training Needs self-administered questionnaire was used to collect data (17). Intellectual property rights for the tool belong to the University of Birmingham (UoB), while the World Health Organization holds the license for online use (17). Therefore, permission to use the tool was not required, but users should acknowledge the UoB and WHO. The tool is psychometrically validated and has been used widely in healthcare. The original questionnaire has 30 items in five categories, namely research/ audit, communication/ teamwork, clinical tasks, administration and supervisory/ management. When adapting the tool, 8 items can be replaced and 10 added without compromising the validity of the instrument (17). For this study, the categories and items included research/ audit category (3, 7, 22 and 25), communication/ teamwork category (1, 5, 6, 11, 12, and 21), clinical tasks category (8, 10, 14, 15, 18, 20, 26, 27, 28, 29, 30, 31, 32, 33, 34, 35, 36 and 37), administration (2, 17 and 23) and supervisory/ management (4, 9, 13, 16, 19 and 25). The adaptation of the tool was done based on a literature review and discussions with experts. Items for MNH were adapted from a study conducted in Tanzania (27). In total, the data collection tool had 37 items.

The original tool has three sections. Section A determines demographic characteristics. Section B asks respondents to rate items across four indicators: perceived task importance (A), perceived task performance (B), if organizational changes could address training need (C) or if the need could be addressed by training courses (D). For this study, only indicators A and B applied. For perceived task importance (A), the importance rating scale used was 1 for “not important” and 7 for “extremely important”. For perceived task performance (B), 1 was rated for “not well” and 7 for “extremely well”.

Originally, Section C has open-ended questions. After pre-testing the tool, the responses were coded based on identified topics. The responses were coded because there was repetition. An option to mention other specific topics was included after the coded items.

### Data collection procedure

Information about the study was communicated to the management of both study sites. Access to registered midwives was obtained via management and in-service coordinators. Information sheets were handed out to eligible midwives. Those willing to participate voluntarily signed the informed consent and were informed of the right to withdraw at any time during the study, without any penalties. Questionnaires were distributed to midwives based on the numbers that had been randomly selected from the random number table. The questionnaire was paper-based and self-administered using a pen. Field workers were on site for assistance with clarity on the questions as they had been trained on the data collection tool. After completion of the questionnaire, they were submitted to the field workers over the course of the day. During night shifts, a sealed box was kept for respondents to drop the questionnaires. Data were collected from 28 November 2023 to 25 January 2024.

### Data analysis

Descriptive statistics were used to determine means and standard deviations for demographic characteristics. The scores for the perceptions of participants were calculated per item in each of the superordinate categories for perceived task importance and perceived performance. To determine training needs, perceived task performance (Indicator B) was subtracted from perceived task importance (Indicator A). Where B was rated lower than A, the difference indicated a training need (17). A previously used criterion of a standard deviation > 1.5 was applied to identify the highest training needs, and the results of a standard deviation <1.5 were not included in the final list (20, 28). A paired sample t-test was performed to determine statistically significant differences between the means of each category pair, and the p-value was p=<. 05.

### Ethics approval

Ethics approval was obtained from the University of Johannesburg Higher Degrees Committee. Approval was also obtained from the University of Johannesburg Research Ethics Committee, protocol number 1966-2023, valid until 22 May 2024 and renewed until 9 August 2025. Ethics clearance was also obtained from Eswatini Health and Human Research Review Board, protocol numbers EHHRRB 086/2023, and EHHRRB 127/2024 after renewal. Permission to gain access to the study sites was obtained from the Ministry of Health of Eswatini for the Hlathikulu Government Hospital for pretesting of the data collection tool and from the two study sites where data were collected. Respondents voluntarily signed the informed consent form before participating in the study. Questionnaires were anonymized with codes and no identifiable data were collected. The ethics principles of beneficence, non-maleficence, justice were observed.

## Results

The mean age of the 202 respondents was 38.06±6.9 years. The midwives predominantly fell into the age group 40–44 years (n=53, 22.2%), and they had an average of 5–9 years of work experience (n=75, 37.1%).) [Table 1].

**Table 1:** Demographic characteristics of respondents.

| Variable | Frequency (n) | Percentage (%) |
| --- | --- | --- |
| *Age in years* |  |  |
| 25–29 years | 21 | 10.4 |
| 30–34 years | 44 | 21.8 |
| 35–39 years | 51 | 25.2 |
| 40–44 years | 53 | 26.2 |
| 45–49 years | 21 | 10.4 |
| 50+ years | 12 | 5.9 |
| *Sex* |  |  |
| Females | 160 | 79.2 |
| Males | 42 | 20.8 |
| *Highest level of education* |  |  |
| Diploma in general nursing with postgraduate diploma certificate in midwifery | 43 | 21.3 |
| Bachelor of nursing with midwifery option | 152 | 75.2 |
| Advanced midwifery | 2 | 1.0 |
| Master’s degree in midwifery | 1 | 0.5 |
| Master’s degree in nursing | 4 | 2.0 |
| *Work experience* |  |  |
| 0–4 years | 71 | 35.1 |
| 5–9 years | 75 | 37.1 |
| 10–14 years | 32 | 15.8 |
| 15–19 years | 14 | 6.9 |
| 20+ years | 10 | 5.0 |
| *Demographic characteristics of study respondents for the two study sites* |  |  |

The training needs of respondents were determined from the mean differences for perceived task importance (Indicator A) and perceived task performance (Indicator B). All 37 items of the questionnaire indicated training needs, as the mean values for Indicator B were lower than means for Indicator A. A previously used criterion was applied to categorize training needs according to highest and lowest training needs (20, 28). The criterion is as follows:

i) All scores < than 1.5 were lower training needs
ii) All scores > 1.5 were higher training needs.

Based on the above criteria, 19 items had high training needs out of the 37 in the questionnaire (S1 Table).

Table 2 shows the highest training needs identified by midwives in the study. The item with the highest training need was critically evaluating research (M=3.02±1.88). The two items with the lowest training needs were using technical equipment, including computers (M=1.68±1.76), and accessing research resources e.g., time, money, information, and equipment (M=1.68± 1.48). The training needs according to the superordinate category pairs are shown below <u>(S1 Fig)</u>.

The results in Figure 1 show the research/ audit category as the highest training need category (2.23±1.05), followed by clinical tasks (1.94±0.55). Respondents also indicated the need for training on administrative tasks (1.70±1.00), communication (1.57±0.79), and supervisory tasks (1.14±0.76).

**Fig 1.**
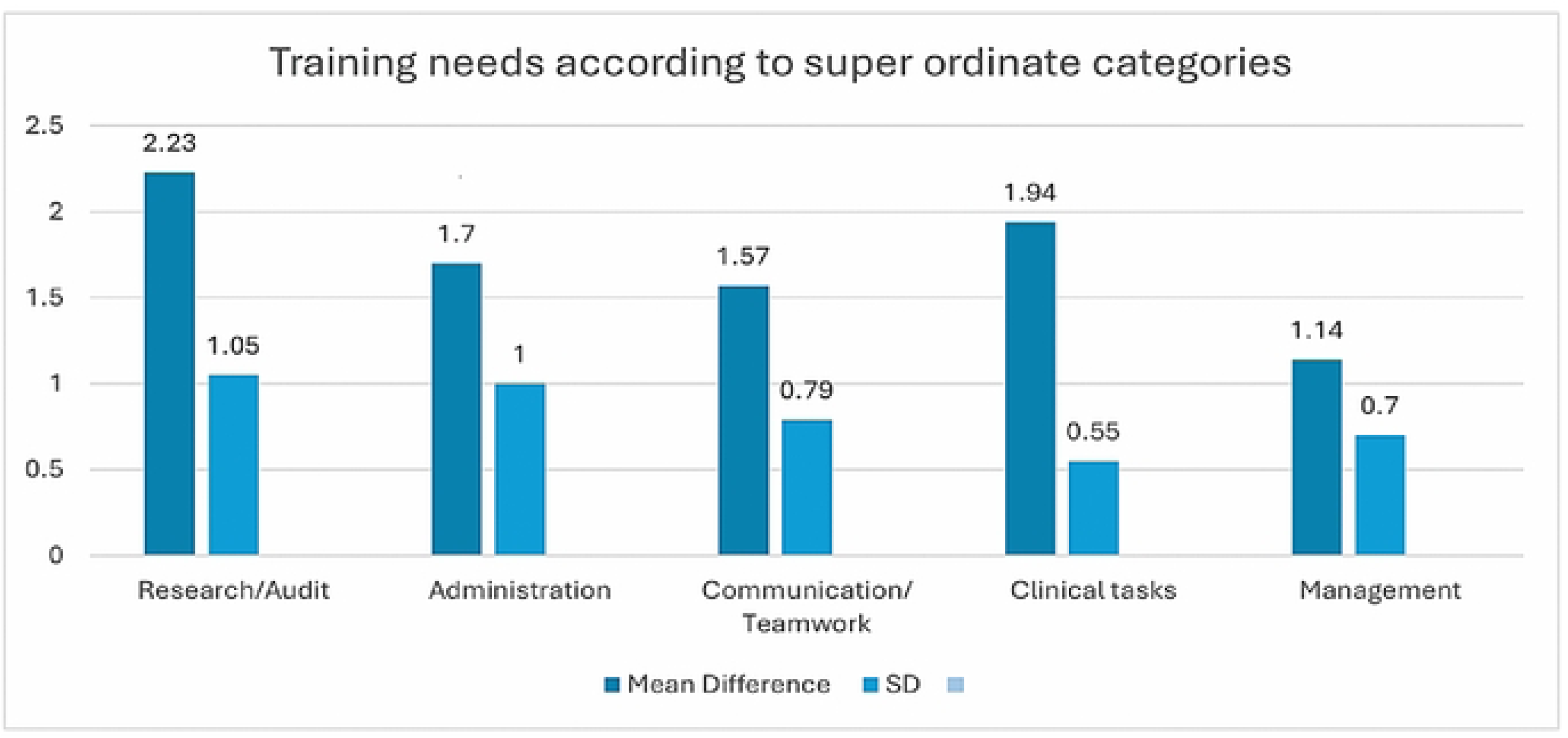
Training needs of registered midwives according to superordinate categories.

The results in Table 3 indicate that there were statistically significant differences between each superordinate category pair on task importance and task performance at p=<.001.

**Table 3.** Paired t-test results comparing the differences in means for each superordinate category pair.

| Superordinate category | TI | TP | Mean Difference | SD | Paired t-test |  |  |  |
| --- | --- | --- | --- | --- | --- | --- | --- | --- |
|  |  |  |  |  | SD EM | t-value | df | Sig (2 tailed) |

|  | M | SD | M | SD |  |  |  |  |  |  |
| --- | --- | --- | --- | --- | --- | --- | --- | --- | --- | --- |
| Research | 5.81 | 0.77 | 3.76 | 0.72 | 2.23 | 1.05 | 0.74 | 26.25 | 201 | <,001* |
| Communication | 6.30 | 0.49 | 4.73 | 0.55 | 1.57 | 0.79 | 0.56 | 28.13 | 201 | <,001* |
| Clinical tasks | 6.66 | 0.40 | 4.95 | 0.34 | 1.94 | 0.55 | 0.39 | 44.49 | 201 | <,001* |
| Administration | 6.05 | 0.67 | 4.41 | 0.66 | 1.70 | 1.00 | 0.71 | 22.50 | 201 | <,001* |
| Supervisory | 6.11 | 0.54 | 4.97 | 0.53 | 1.14 | 0.76 | 0.54 | 21.32 | 201 | <,001* |

*Differences in means between the five category pairs*

Table 4 shows the specific training needs for registered midwives from both study sites. In total, seven specific needs were identified form the coded section of the tool. The highest specific need was newborn resuscitation, while the lowest was the management of preeclampsia and eclampsia, and the management of critically ill newborns.

**Table 4.**
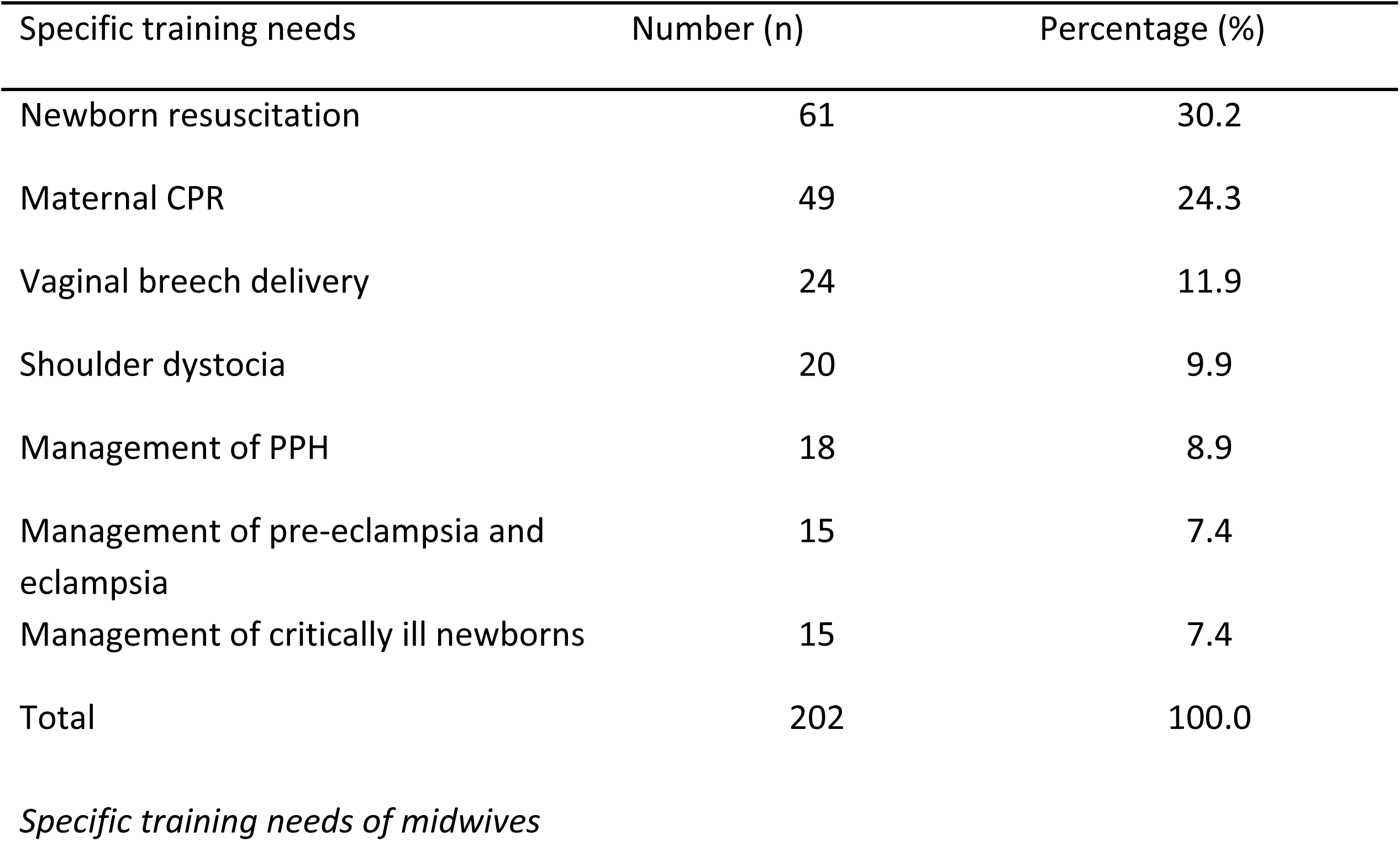
Specific training needs for registered midwives.

In the open-ended option, 14 specific training needs were mentioned. The respondents repeated newborn resuscitation (n=48, 23.8%), the management of preeclampsia and eclampsia and PPH (n=16, 7.9%). Other mentioned topics for training included the management of puerperal sepsis (n=32, 15.8%), obstructed labor (n=21, 10.4%), the management of APH (n=17, 8.4%), respectful maternity care (n=11, 5.4%), ongoing EmONC training (n=9, 4.5%), CPAP (n=7, 3.5%), preterm care and vacuum delivery (n=6, 3.0% each), the manual removal of retained placenta (n=5, 2.5%), ongoing training on the use of critical care equipment and early identification of danger signs in pregnancy (n=4, 2.0% each).

## Discussion

The aim of the study was to determine the training needs of midwives from two CEmONC facilities to inform a training program.

The results from the study reveal that all the 37 items measured in the questionnaire indicated training needs. The findings are consistent with a study on the training needs of healthcare professionals in Greece (29) and in Nigeria (30). The results suggest that midwives require training across all the items measured in the questionnaire. Nineteen items could be identified as the highest training needs. These related mostly to clinical tasks. In another study, the training needs of oncology professionals were also clinical tasks (22). The clinical work of midwives who work at CEmONC facilities could have influenced the high rating on clinical tasks.

Training needs according to the five categories showed that the research/ audit category had the highest training need. The finding on the research category is consistent with several TNA studies (19, 20, 23). Conversely, clinical tasks (31) and communication (30) were identified as high training needs. The results show varied training needs across different study populations (30).

All of the items in the research category were indicated as high training needs. The research ratings could be due to inadequate preparation for research use during nursing and midwifery education (32). Yet, midwives have to be able to conduct research and use it in clinical practice to provide evidence-based maternity care (33). Research interest could develop through training, collaboration, and access to research funds (34).

The clinical tasks category was rated second in the study. Similar findings were noted in a study of primary care nurses in South Africa (19) and among healthcare professionals in Eswatini (18). Low ratings on the perceived task performance of clinical skills implies deficiencies in skills related to the clinical tasks that were measured in the questionnaire. Similar ratings on the clinical task items related to treating patients and assessing patients’ clinical needs were reported in a similar TNA (30). The ratings could be related to the knowledge and skills gaps identified during the audits of perinatal deaths in the period 2014–2016 (35). The training needs with respect to clinical tasks provide a baseline of the skills that should be strengthened for the improved management of obstetric and newborn complications.

Respondents rated the administrative category third. The administration category was also rated third in a study of clinical nurses in Ibadan (30). Two items were rated high in this category, undertaking administrative services and using technical equipment, including computers. The findings are similar to a study on the training needs of healthcare professionals in Sri Lanka (20). In the study setting, the need for training in administrative skills is tied to the fact that midwives participate in administrative tasks in hospitals as they are more senior than staff nurses and nurse assistants. Using technical equipment has been identified as a training need in other studies (19, 36). The use of technology requires competence to minimize errors and regular education is necessary to improve competencies (37).

Fourth in the ratings was the communication category. The items establishing a relationship with patients, getting on with your colleagues, and working as a member of a team were identified as training needs. On the first item, (19, 30) had similar findings. This finding varied from other training needs analysis studies where communication was rated third (20) and last (19, 23). Communication is vital for efficiency and effectiveness during the management of emergency complications. Interprofessional collaboration, hierarchy, and conflicts that emanate from professional boundaries influence communication among physicians, midwives and nurses (38). Communication training is necessary to address communication barriers (39). The training also improves communication among healthcare professionals, clients and their partners, thus improving patient safety (40).

Respondents rated the management category last, especially on the item of introducing new ideas at the workplace. The finding is consistent with those of Adewole, Salawu (30). The training need could be influenced by external stimuli that bring change in healthcare environments (41). Emergency obstetrics management requires the use of current evidence-based care, including EmONC procedures. These need skills for introducing and managing change. Midwives assume more management tasks in the clinical area than other nursing cadres, creating the perceived need.

### Specific training needs

Specific training needs related to midwifery clinical tasks. Similarly, in a study of primary healthcare nurses, clinical tasks were identified (19). Similarities and variances in specific training needs have been noted in literature across study populations and contexts. The clinical needs of respondents could have influenced the identification of the specific training needs. In this study, specific training needs related to management of the leading causes of maternal and newborn mortalities, ongoing training, and respectful maternity care. Early identification of danger signs in pregnancy was also mentioned.

## Conclusion

The purpose of the study was to determine the training needs of registered midwives to inform a training program. Based on the training needs results, training programs should be tailored to training on research, clinical, administrative, and communication tasks. These tasks expand the functioning of the midwife within the professional framework and the scope of midwifery practice. Integrating the identified tasks into continuing professional development activities would channel training resources where required, especially because training is costly and has to be worthwhile. Health outcomes would also improve once there is competent practice after training.

## Limitations and recommendations

The use of a self-reported questionnaire could have led to recall bias, which would influence ratings in the study. Data were collected from two CEmONC hospitals in two cities, excluding rural and private hospitals and BEmONC facilities. There might be slight differences in the obstetric complications in other geographic regions, which would bring about differences in training needs. Despite the limitations, the results of the study provide information on training needs at two facilities that have had persistent adverse health outcomes that should be addressed. A recommendation for the Ministry of Health is the allocation of resources for the training of midwives on all the identified training needs, including research, administration and communication to improve competent practice and multi-professional team collaboration during obstetric emergency events.

## Data Availability

All data files are available from the DRYAD database (DOI:10.5061/dryad.kh18932p5).

## Data and materials

The data for the study are attached.

## Acknowledgments

I would like to acknowledge the immense contribution of my supervisor, Professor R. Ngunyulu, towards the accomplishment of this work. I am also grateful to the Eswatini Ministry of Health and the Eswatini Human and Health Research Review Board for allowing me to conduct the study in the study sites. Appreciation is also extended to the management and in-service coordinators of Hlatikhulu Government Hospital, Mbabane Government Hospital and Raleigh Fitkin Memorial Hospital for allowing me to collect data from the registered midwives in these facilities. I pass my sincere gratitude to the midwives who participated in the study.

## Supporting information

<u>S1 Table.</u> **The training needs of registered midwives from the two study sites**

https://

(DOCX)

<u>S1 Figure.</u> **Training needs according to superordinate categories**

https://

(DOCX)

## References

1. WHO. World Health Organization 2025 data, Eswatini Online: WHO; 2025 [updated 2024; cited 2025 10 July]. Available from: https://data.who.int/countries/748.

2. UN. The sustainable development goals report 2025. United nations, 2025 20 September 2025. Report No. 10. Available from: https://unstats.un.org/sdgs/report/2025/The-Sustainable-Development-Goals-Report-2025.pdf

3. CSO. The 2017 population and housing census The Kingdom of Eswatini Ministry of Health,, Statistics; 2019 08 December 2024. Report No. Volume 3. Available from: https://www.gov.sz/images/FinanceDocuments/Volume-3.pdf

4. CSO. Eswatini Multiple Indicator Cluster Survey 2021-2022, Survey Findings Report,. Government of the Kingdom of Eswatini, 2024 19 June 2025. Report No. 6. Available from: https://www.unicef.org/eswatini/media/1891/file/Eswatini_MICS_Report_2024.pdf.pdf

5. WHO. Trends in maternal mortality 2000 to 2023: estimates by WHO, UNICEF, UNFPA, World Bank Group, and UNDESA/Population Division. Geneva: World Health Organization, 2025 13 August 2025. Report No. Available from: https://www.who.int/publications/i/item/9789240068759

6. Cresswell JA, Alexander M, Chong MYC, Link HM, Pejchinovska M, Gazeley U, et al. Global and regional causes of maternal deaths 2009-20: a WHO systematic analysis. The Lancet Global Health. 2025;13(4):e626–e34.

7. van den Broek N. Keep it simple – Effective training in obstetrics for low- and middle-income countries. Best Practice & Research Clinical Obstetrics & Gynaecology. 2022;80:25–38.

8. Souza JP, Day LT, Rezende-Gomes AC, Zhang J, Mori R, Baguiya A, et al. A global analysis of the determinants of maternal health and transitions in maternal mortality. The Lancet Global Health. 2024;12(2):e306–e16.

9. WHO, UNFPA, UNICEF, AMDD. Monitoring emergency obstetric care: a handbook. WHO Press, 2009 13 July 2021. Report No.: 978 92 4 154773 4. Available from: https://iris.who.int/bitstream/handle/10665/44121/9789241547734_eng.pdf?sequence=1

10. ICM. ICM Essential Competencies for Midwifery Practice. The Hague: ICM, 2024 25 January 2025. Report No. Available from: https://internationalmidwives.org/resources/essential-competencies-for-midwifery-practice/

11. Nove A, Friberg IK, de Bernis L, McConville F, Moran AC, Najjemba M, et al. Potential impact of midwives in preventing and reducing maternal and neonatal mortality and stillbirths: a Lives Saved Tool modelling study. The Lancet Global Health. 2021;9(1):e24–e32.

12. ICM. Building the enabling environment for midwives: A call to actions for policy makers. The Hague: ICM, 2021 18 May 2024. Report No. Available from: https://www.internationalmidwives.org/wp-content/uploads/11061-eng_icm-enabling-environment-policy-brief_v1.1_20210629.pdf

13. Woodman S, Bearman C, Hayes P. Understanding skill decay and skill maintenance in first responders. The Australian Journal of Emergency Management. 2021;36(4):44–9.

14. Klostermann M, Conein S, Felkl T, Kluge A. Factors influencing attenuating skill decay in high-risk industries: A scoping review. Safety. 2022;8(2):1–23.

15. Puspita S, Nurhalim A. IMPORTANCE OF TRAINING NEEDS ANALYSIS FOR HUMAN RESOURCES DEVELOPMENT IN ORGANIZATIONS. E-Mabis: Jurnal Ekonomi Manajemen dan Bisnis. 2021;22:151–60.

16. Markaki A, Malhotra S, Billings R, Theus L. Training needs assessment: Tool utilization and global impact. Bmc Medical Education. 2021;21(1):1–20.

17. Hennessy DA, Hicks CM. Training needs analysis questionnaire and manual. University of Birmingham. 2011. Available from: http://www.who.int/workforcealliance/knowledge/HennessyHicks_trainingneedstool.pdf.

18. Steinhorst J, Baker C, Padidar S, Litschka-Koen T, Ngwenya E, Mmema L, et al. Developing and applying a TNA tool for healthcare workers managing snakebite envenoming: A cross-sectional study in Eswatini. Plos Neglected Tropical Diseases. 2025;19(1):e0012778.

19. Kordom A, Daniels F, Chipps J. Training needs of professional nurses in primary health care in the Cape Metropole, South Africa. African journal of primary health care & family medicine. 2023;15(1):e1–e10.

20. Rajapakshe W, Weerarathn RS, Pathirana GY, Malage MH. Analysis on current and future training needs in health sector of Sri Lanka. Calitatea. 2022;23(189):277–88.

21. Adewole DA, Negedu NS, Bello S. Preferred approach to clinical performance improvement among physicians at the university college hospital, Ibadan Nigeria. Nigerian Journal of Medicine. 2021;30:314–9.

22. Byamugisha J, Munabi IG, Mubuuke AG, Mwaka AD, Kagawa M, Okullo I, et al. A health care professionals training needs assessment for oncology in Uganda. Human Resources for Health. 2020;18(1):1–26.

23. Adewole DA, Adeniji FIP, Makanjuola AV. Self-Reported Training Needs among Physicians in a Tertiary Institution, Southwest, Nigeria: An Application of Hennessy-Hicks Training Needs Assessment Tool. Nigerian Journal of Medicine. 2020;29(3):396–400.

24. Pavlidis G, Downs C, Kalinowski TB, Swiatek-Barylska I, Lazuras L, Ypsilanti A, et al. A survey on the training needs of caregivers in five European countries. Journal of nursing management. 2020;28(2):385–98.

25. Magagula SV. A case study of the Swaziland Essential Health Care Package; Discussion paper 112. Harare: MoH Swaziland, 2017.

26. bin Ahmad H, binti Halim H. Determining sample size for research activities: The case of organizational research. Selangor Business Review. 2017;2(1):20–34.

27. Mwansisya T, Mbekenga C, Isangula K, Mwasha L, Pallangyo E, Edwards G, et al. Translation and validation of TNA questionnaire among reproductive, maternal and newborn health workers in Tanzania. BMC Health Services Research. 2021;21(1):1–12.

28. Shongwe NG. LEARNING NEEDS ASSESSMENT FOR CONTINUOUS PROFESSIONAL DEVELOPMENT IN PAEDIATRIC ACUTE CARE NURSING: THE ESWATINI PROJECT. Master’s dissertation, University of Witwatersrand; 2019.

29. Tsantili I, Hadjidema S, Galanis P. Identification of the Training Needs of Health Care Professionals in Greece. International Journal of Caring Sciences. 2021;14(1):115–26.

30. Adewole DA, Salawu MM, Bello S. Training needs assessment and preferred approach to enhancing work performance among clinical nurses in University College Hospital (UCH), Ibadan, Oyo State, South-western Nigeria. International Journal of Nursing and Midwifery. 2020;12(4):130–8.

31. Holloway K, Arcus K, Orsborn G. TNA – The essential first step for continuing professional development design. Nurse Education in Practice. 2018;28:7–12.

32. Owusu LB, Scheepers N, Tenza IS. Exploring nurse educators’ preparation of clinical nurses and midwives for research utilization in practice: A qualitative study. Nurse Education Today. 2025;145:106476.

33. Shadap A. Evidence based practice in midwifery care. International Journal of Obstetrics and Gynaecological Nursing. 2022;4:1–4.

34. Kyei J, Dzansi G, Acheampong AK, Adjei CA, Ohene LA, Adjorlolo S, et al. Factors Influencing Nurses and Midwives’ Participation in Research: A Qualitative Study. Nursing & Midwifery Research Journal. 2023;19(1):5–21.

35. Government of the Kingdom of Swaziland. Confidential enquiry on maternal deaths:Triennial Report 2014-2016. Ministry of Health, 2017 07 January 2022. Report No.

36. Burke D, Cocoman A. Training needs analysis of nurses caring for individuals an intellectual disability and or autism spectrum disorder in a forensic service. Journal of Intellectual Disabilities and Offending Behaviour. 2019;11(1):9–22.

37. Konttila J, Siira H, Kyngäs H, Lahtinen M, Elo S, Kääriäinen M, et al. Healthcare professionals’ competence in digitalisation: A systematic review. Journal of Clinical Nursing. 2019;28(5-6):745–61.

38. Schmiedhofer M, Derksen C. Barriers and facilitators of safe communication in bstetrics: Results from qualitative interviews with physicians, midwives and nurses. Int J Environ Res Public Health. 2021;18(3):1–16.

39. Hüner B, Derksen C, Schmiedhofer M, Lippke S, Riedmüller S, Janni W, et al. Reducing preventable adverse events in obstetrics by improving interprofessional communication skills – Results of an intervention study. BMC Pregnancy and Childbirth. 2023;23(1):1–13.

40. Lippke S, Derksen C, Keller F, Kötting L, Schmiedhofer M, Welp A. Effectiveness of communication interventions in obstetrics—A systematic review. International journal of environmental research and public health. 2021;18:2616.

41. Milella F, Minelli EA. Change and Innovation in Healthcare: Findings from Literature. 2021;13:395–408.

